# BLOOD-BASED BIOMARKERS TO IDENTIFY AND MONITOR RECURRENT ENDOMETRIAL CANCER: A SYSTEMATIC REVIEW AND META-ANALYSIS

**DOI:** 10.1101/2025.08.28.25333862

**Authors:** Anisha Chitrakar, Ayisha A Ashmore, Sylwia Bujkiewicz, Lorna Wheaton, Coral Pepper, David S Guttery, Esther L Moss

## Abstract

**Objectives:** The detection of endometrial cancer recurrence is primarily through patient reported symptoms and can occur many years after primary treatment. Investigation of suspected endometrial cancer recurrence requires clinical examination, imaging, and if a lesion is identified a biopsy for pathological analysis. A blood-based biomarker could support the identification and confirmation of endometrial cancer recurrence.

**Methods:** A systematic review was undertaken to identify studies that reported the use of biomarkers in the monitoring and/or diagnosis of endometrial cancer recurrence (PROSPERO registration CRD42021226204). MEDLINE, Embase, Emcare, CINAHL, the Cochrane Central Register of Controlled Trials (CENTRAL), and OpenGrey were searched up to 15^th^ November 2024. Studies were assessed using QUADAS-2. Meta-analysis of sensitivity and specificity was carried out using Bayesian random effects univariate meta-analytic models for sensitivity and specificity individually.

**Results:** Of the 10,643 references identified, 142 studies underwent full-text assessment and data was extracted from 25 studies. CA125, HE4, circulating free or tumour DNA (cf/ctDNA), and carcinoembryonic antigen (CEA) were the most extensively studied biomarkers. Although ten studies investigated the use of more than one biomarker, the majority were comparing rather than combining their diagnostic ability. Sensitivity was highest for cf/ctDNA, 0.87 (95% CrI: 0.63, 0.99), followed by HE4, 0.74 (95% CrI 0.51-0.90). Specificity was highest for CA125; 0.91 (95% CrI: 0.77, 0.99) followed by cf/ctDNA 0.89 (95% CrI: 0.63-0.99).

**Conclusions:** cf/ctDNA had had superior diagnostic accuracy to detect endometrial cancer recurrence compared to CA125 and HE4. Given the concerning rise in endometrial cancer mortality, future research should focus on a potential role for a blood-based biomarker in facilitating the identification of endometrial cancer recurrence and the impact on survival following recurrence.

**KEY MESSAGES:** *What is already known on this topic:* Blood-based biomarkers have the potential to monitor and/or detect endometrial cancer recurrence.

*What this study adds:* circulating free/tumour DNA had had superior diagnostic accuracy to detect endometrial cancer recurrence compared to other blood-based biomarkers for example CA125 and HE4

*How this study might affect research, practice or policy:* the results indicate a potential role for circulating free/tumour DNA in detecting endometrial cancer.

## INTRODUCTION

Endometrial cancer is typically viewed as a malignancy associated with a good long-term outcome, with 72% of women diagnosed in the UK surviving for more than 10 years following diagnosis ^(1)^, and the majority of patients dying of non-endometrial cancer associated causes ^(2)^. However, recent data showing rising mortality rates ^(3)^, particularly amongst women from ethnic minority groups ^(4)^, is challenging this view. As a result, there are increasing calls for the management of endometrial cancer recurrence to undergo the same paradigm shift that has taken place with recurrent ovarian cancer, where survival benefits have been reported with secondary cytoreductive surgery ^(5)^. Indeed, the survival benefits from surgery in selected endometrial cancer recurrence cases can be considerable, however prognostic factors reported to impact the success of salvage treatment are tumour size ^(6)^ and the time to recurrence ^(7, 8)^.

The detection of endometrial cancer recurrence is primarily through patient reported symptoms, and although intensive follow-up has not been shown to improve overall survival ^(9)^, symptomatic presentation is reportedly associated with lower treatment rates and worse survival compared to those who were asymptomatic at recurrence diagnosis, 6.5 months versus 11.1 months, p=0.0053 ^(10)^. The introduction of molecular classification of endometrial cancer has led to prognostication and risk stratification ^(11)^, however the lack of a sensitive and specific marker that can be used to detect, or predict endometrial cancer recurrence can make identifying patients at risk of recurrence more challenging, and can lead to increased use of imaging or biopsies if recurrence is suspected. The use of the blood-based ovarian cancer tumour markers CA125 and HE4 has been reported ^(12)^, however the developments in liquid-based technology have opened new options for the detection of endometrial cancer recurrence^(13)^.

The aim of this systematic review was to investigate the diagnostic accuracy of blood-based biomarkers to monitor and/or detect endometrial cancer recurrence, and if sufficient data available to perform a meta-analysis comparing the sensitivity/specificity.

## METHODS

The protocol for the systematic review was registered with PROSPERO on 03/02/2021 (CRD42021226204). Transparent Reporting of Systematic Reviews and Meta-analyses (PRISMA) Statement guidelines^15^ were followed and checklist completed. All studies were independently reviewed by two reviewers and conflicts were resolved through discussion with a third reviewer. The search aimed to identify studies that reported the use of biomarkers in the monitoring and/or diagnosis of endometrial cancer recurrence.

### Information sources and search strategies

Searches were conducted by a clinical librarian. MEDLINE, Embase, Emcare, CINAHL, the Cochrane Central Register of Controlled Trials (CENTRAL), and OpenGrey were searched on the 3rd June 2020 and updated 15^th^ November 2024. The strategies were developed using a range of keywords and thesaurus terms in the structure: endometrial cancer AND biomarkers AND diagnostic studies. The diagnostic studies section was adapted from the OVID diagnostic studies filter (https://tools.ovid.com/ovidtools/expertsearches.html). A human filter was applied in the strategy, but no date or language limits were utilised. The detailed search description is available in Supplementary Data 1.

### Study eligibility criteria

Identified studies were uploaded to the Covidence software (Melbourne, Victoria, Australia), a web-based systematic review program. Duplicates were removed and the remaining studies screened by title and abstract. The eligibility criteria included: histological diagnosis of endometrial cancer; minimum of 3 months since completing primary treatment for endometrial cancer; minimum of 5 patients; blood-based biomarker; including cases of endometrial cancer recurrence; and peer-reviewed literature. Studies containing ovarian cancer and cervical cancer patients where the data from endometrial cancer patients could not be separated were excluded, as were studies focused on uterine sarcoma or endometrial stromal neoplasms, and non-human studies.

### Data extraction and quality assessment

Data extraction was performed using the Covidence software and included study year, population, histological subtype, biomarker thresholds, test accuracy for detection of recurrence (sensitivity, specificity, positive predictive value (PPV), negative predictive value (NPV) and area under the curve (AUC) value). The revised Quality Assessment of Diagnostic Accuracy Studies (QUADAS-2) was used to evaluate the quality of the studies included. Each study was assessed by two reviewers independently and conflicts/uncertainties were discussed with a third reviewer, in order to gain resolution. Data was summarised from the identified studies including patient demographics, histological subtype (where mentioned), duration of follow-up, and the number of recurrences. In accordance with the journal’s guidelines, we will provide our data for independent analysis by a selected team by the Editorial Team for the purposes of additional data analysis or for the reproducibility of this study in other centers if such is requested.

### Statistical methods

Meta-analysis of sensitivity and specificity was carried out using random effects univariate meta-analytic models for sensitivity and specificity individually. Sensitivity analysis using the fixed effects model was carried out for data sets where data from no more than three studies were available. The meta-analyses were implemented in a Bayesian framework using WinBUGS14 software. Additional analyses and visualisations were carried out using RStudio. The results are presented as means and 95% credible intervals (CrIs).

## RESULTS

A total of 14,850 references were retrieved from different databases, 4,207 duplicate entries were removed, and 10,643 unique studies were selected for title and abstract screening. During this process, 10,501 studies were excluded as they did not align with the predefined inclusion criteria due to unrelated cancer types, irrelevant outcomes, or non-human subjects. The remaining 142 were then subjected to an intense full-text review. At this stage, 117 studies were excluded for not meeting the study eligibility criteria. Commonly cited reasons for exclusions included studies with patient sampling only at the time of diagnosis, no longitudinal data on recurrence, and inability to separately identify endometrial cancer patients from patients with other cancer primaries. Ultimately, a total of 25 studies fulfilled all the inclusion criteria for this review and examined various blood biomarkers (Figure 1). CA125 ^(12, 14–27)^, HE4 ^(12, 20–25)^, circulating free or circulating tumour DNA (cf/ctDNA) ^(25, 28–30)^, and carcinoembryonic antigen (CEA) ^(31–34)^ were the most extensively studied biomarkers (Table 1). In 10 studies more than one biomarker was investigated ^(12, 18, 20–25, 31, 34)^.

**Figure 1.**
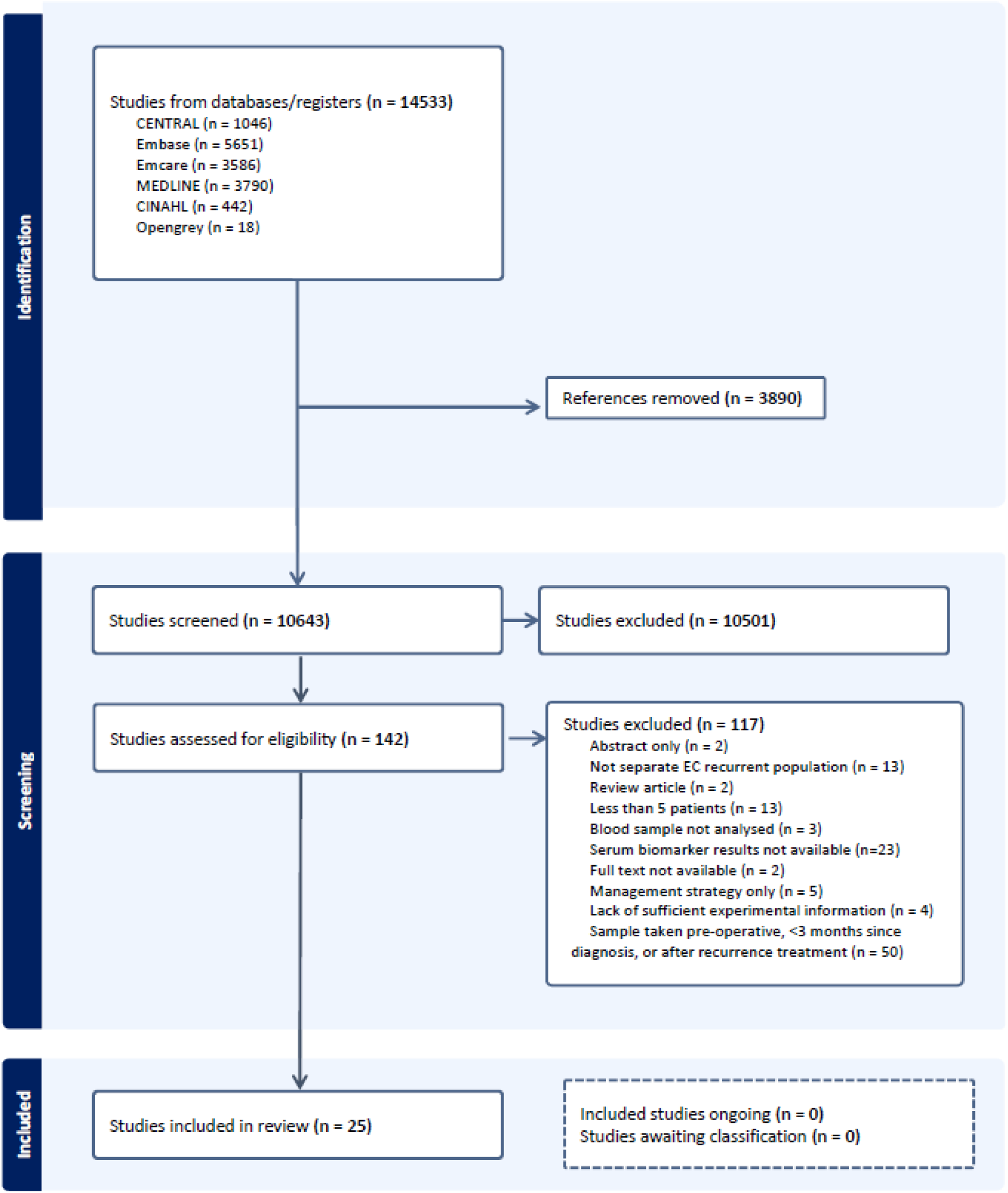
PRISMA flow chart.

**Table 1.** Summary of the studies included. E= endometrioid histology; NonE = non-endometrioid histology; AC = adenocarcinoma; Y = years; R = recurrence; NR = no recurrence; ^†^calculated from supplementary data. ^$^ 41 recurrences reported within the cohort however only 38 had a CA125 result.

| Year | Author | Cases | Biomarker | Median age (range) | Histology | Follow-up months (range) | Recurrences (percentage) |
| --- | --- | --- | --- | --- | --- | --- | --- |
| 1985 | Paulick <sup>(31)</sup> | 206 | TPA, PHI, CEA | Unknown | Unknown | Unknown | 31 (15%) |
| 1990 | Patsner <sup>(14)</sup> | 125 | CA125 | Unknown | E: 121<br>NonE: 4 | 18 (12-36) | 13 (10.4%) |
| 1991 | Fanning <sup>(15)</sup> | 21 | CA125 | Unknown | Unknown | 12 (9-38) | 9 (42.8%) |
| 1994 | Rose <sup>(16)</sup> | 266 | CA125 | Unknown | Unknown | 39 (4-54) | 33 (12.4%) |
| 1998 | Price <sup>(17)</sup> | 15 | CA125 | Unknown | NonE: 15 | 63 (21-90) | 5 (33%) |
| 1999 | Lo <sup>(18)</sup> | 23 | CA19.9, CA125, CA15.3 | Unknown | Unknown | Unknown | 7 (30.4%) |
| 2003 | Bruns <sup>(32)</sup> | 126 | CEA | (18-85) | AC: 122<br>NonE: 6 | 62.1 (2.6-161.4) | 10 (7.8%) |
| 2013 | Frimer <sup>(19)</sup> | 56 | CA125 | R: mean 68.0<br>NR: mean 65.4 | NonE: 56 | Unknown | 23 (41%) |
| 2013 | Gong <sup>(20)</sup> | 434 | HE4, CA125 | 52.5 (24-81) | E: 434 | 53.26 (6-111) | 37 (8.5%) |
| 2015 | Angioli <sup>(21)</sup> | 252 | HE4, CA125 | mean 61.2 (38-81) | E: 228<br>NonE: 24 | 38 (24-75) | 37 (14.7%) |
| 2015 | Brennan <sup>(22)</sup> | 98 | HE4, CA125 | 65 (40-85) | E: 69<br>NonE: 30 | 5 years (0.6-12.6Y) | 26 (26.5%) |
| 2015 | Hashiguchi <sup>(35)</sup> | 146 | Sialyl | 60 (28-83) | E: 132<br>NonE: 14 | 44 (1-83) | 29 (19.9%) |
| 2015 | Kozakiewicz <sup>(36)</sup> | 317 | TATI | mean 60.5 (32-81) | AC: 244<br>NonE: 73 | max 16 years | 59 (18.6%) |
| 2016 | Hashiguchi<sup>(33)</sup> | 215 | CEA | 60 (28-85) | E: 191<br>NonE: 24 | 45<br>(1-95) | 52<br>(24.2%) |
| 2016 | Kozakiewicz<sup>(34)</sup> | 316 | TATI,<br>CEA | 61 (32-81) | Unknown | 2.98 years<br>(0.24-16.93Y) | 59<br>(17.9%) |
| 2016 | Lemech<sup>(37)</sup> | 30 | CTCs | mean 66<br>(49-83) | Unknown | Unknown | 17<br>(56.7%) |
| 2018 | Abbink<sup>(23)</sup> | 157 | HE4,<br>CA125 | 63<br>(56-71) | E: 122<br>NonE: 35 | max 5 years | 48<br>(30.5%) |
| 2020 | Moss<sup>(28)</sup> | 13 | ctDNA | Unknown | E: 8<br>NonE: 5 | Unknown | 9<br>(69.3%) |
| 2021 | Cymbaluk-Ploska<sup>(24)</sup> | 349 | HE4,<br>CA125 | mean 60.8<br>(36-79) | E: 302<br>NonE: 47 | max 52 | 101<br>(28.9%) |
| 2021 | Feng<sup>(25)</sup> | 9 | ctDNA,<br>CA125,<br>HE4 | (42-75) | E: 1<br>NonE: 8 | Unknown | 3<br>(33%) |
| 2021 | Quan<sup>(12)</sup> | 191 | HE4,<br>CA125 | (35-89) | E: 70<br>NonE: 121 | 27<br>(2-94) | 42<br>(22%) |
| 2023 | Ashley<sup>(29)</sup> | 44 | cfDNA | Unknown | E: 24<br>NonE: 20 | 33<br>(0.3-44) | 11<br>(25%) |
| 2023 | Li<sup>(26)</sup> | 1739 | CA125 | Unknown | E: 1643<br>NonE: 96 | Unknown | 120<br>(7.4%) |
| 2023 | Wu<sup>(27)</sup> | 518 | CA125 | 57<br>(22-87) | E: 465<br>NonE: 53 | 49<br>(7-136) | 41<sup>s</sup><br>(7.9%) |
| 2024 | Recio<sup>(30)</sup> | 101 | ctDNA | 67<br>(32-88) | E: 37<br>NonE: 61 | 6.8<br>(0.37-19.1) | 8<sup>†</sup><br>(7.9%) |

**Table 2.** Heterogeneity parameters tau with 95% credible interval (CrIs), Ns - number of studies for each analysis.

|  | sensitivity |  | specificity |  |
| --- | --- | --- | --- | --- |
| Biomarker | Ns | tau (95% CrI) | Ns | tau (95% CrI) |
| CA125 | 10 | 1.15 (0.52, 1.9) | 9 | 1.65 (1.11, 1.99) |
| HE4 | 3 | 0.61 (0.02, 1.8) | 4 | 1.20 (0.56, 1.94) |
| CEA | 3 | 0.60 (0.02, 1.8) | 2 | 1.02 (0.12, 1.94) |
| ctDNA | 4 | 1.20 (0.12, 1.96) | 4 | 0.92 (0.04, 1.93) |

### Quality assessment

The quality assessment of studies included risk of bias: patient selection, index test, reference standard and flow/timing; and applicability concerns: patient selection and reference standard (Supplementary data 2). The majority of studies were real-world single-gate studies and the method of diagnosis of recurrence was not recorded. For several studies the number of participants who had an abnormal biomarker result was not given, only the pooled value in the recurrence population, for example ^(21, 23)^.

### Characteristics of studies and populations

The 25 identified studies showed marked heterogeneity in design, demographics assessed and information provided. Sample sizes varied, from as few as nine ^(25)^ to 1739 ^(26)^ participants, although the majority of studies contained between 100 to 300 participants. The age distribution of the patients was wide, ranging from 30 to 80 years of age, representing the full breadth of the endometrial cancer population. The largest histological subgroup was endometrioid, reflecting the endometrial cancer population, although two studies focused solely on serous histology cases ^(17, 19)^. None of the studies categorised their cases using the TCGA or PROMISE classification, a reflection that the majority of studies were conducted prior to the adoption of molecular classification. Location of recurrence was not mentioned consistently, however some studies noted that location was associated with biomarker positivity, for example none of the pelvic recurrences had a positive CA125 test ^(14)^.

### CA125

CA125 received the greatest focus, 15 studies either alone or combined with another biomarker. The standard cut off of 35 U/mL was used, although in one study a threshold of 26.4 U/mL was used ^(20)^, in another 32.45 U/mL ^(12)^, and Wu et al., ^(27)^ included two thresholds, 35 and 13.75 U/mL. Information on sensitivity was available from 10 studies ^(14–19, 22, 23, 25, 27)^ and specificity from 9 studies ^(14, 15, 17–19, 22, 24, 25, 27)^.

### HE4

HE4 was the focus of seven studies, all of which compared its use with CA125. The test threshold 70 pmol/L was used in all of the studies; however, this threshold was only used in the premenopausal population in three studies ^(12, 20, 25)^, and a threshold of 140 pmol/L was used instead for the postmenopausal population ^(12)^, or over 70-year-old participants ^(25)^. In addition, three studies analysed the data using non-standard thresholds 45.5 pmol/L ^(20)^, 92.5 pmol/L ^(12)^ and 201.3 pmol/L ^(21)^. Information on sensitivity was available in three studies ^(21–23)^ and specificity in four studies ^(21, 22, 24, 25)^.

### Circulating free/tumour DNA

The use of cf/ctDNA to detect recurrence was reported in four studies ^(25, 28–30)^ although the number of participants was lower, with three out of four studies containing less than 50 participants. The studies focused predominantly on the genomic analysis of cf/ctDNA, however sensitivity/specificity for all four studies could be determined from data within the papers or supplementary data, although a time point of 5 months rather than 3 months had to be taken with one study [40].

### CEA

CEA was the focus of four studies, three of which used the threshold of 5 ng/mL ^(31–33)^. Kozakiewicz et al ^(34)^ reported a sensitivity of 76% and specificity of 54.3%, much higher than that calculated for the other three studies, but in light of no test threshold being given it could not be included in the meta-analysis. Three studies were included in the sensitivity ^(31–33)^ and two in the specificity analysis ^(31, 33)^.

### Other biomarkers and combinations

Several less commonly studied biomarkers were included in the review. Sialyl-Tn levels did appear to reflect disease activity ^(35)^, whereas TATI was associated with high sensitivity but low specificity thereby limiting its utility as an independent biomarker marker ^(36)^. Circulating tumour cells, as with cf/ctDNA, demonstrated their potential to predict prognosis, with their presence being associated with shorter recurrence-free survival ^(37)^.

Although ten studies investigated the use of more than one biomarker, the majority were comparing rather than combining their diagnostic ability. Quan et al., ^(12)^ reported that the combination of CA125 and HE4 led to higher sensitivity and specificity than either biomarker separately. Similarly, Feng et al., ^(25)^ reported that integrating cfDNA with other biomarkers (CA125 and HE4) improved predictive accuracy.

### Meta-analysis

A meta-analysis of sensitivity (Figure 2) and specificity (Figure 3) was performed with the available data. The estimates for the individual studies represent the model estimates from the random effects meta-analysis. cf/ctDNA was the most sensitive endometrial cancer marker at 0.87 (95% CrI: 0.63, 0.99); whereas CA125 was the most specific; 0.91 (95% CrI: 0.77, 0.97) compared to 0.89 (95% CrI: 0.70-0.98) for cf/ctDNA. Between-study heterogeneity parameters were estimated with large uncertainty for data sets within a small number of studies. Sensitivity analysis using fixed-effects model yielded sensitivity of 0.74 (0.64, 0.82) for HE4 and 0.26 (0.18, 0.35) for CEA, and specificity of 0.89 (0.85, 0.93) for CEA.

**Figure 2.**
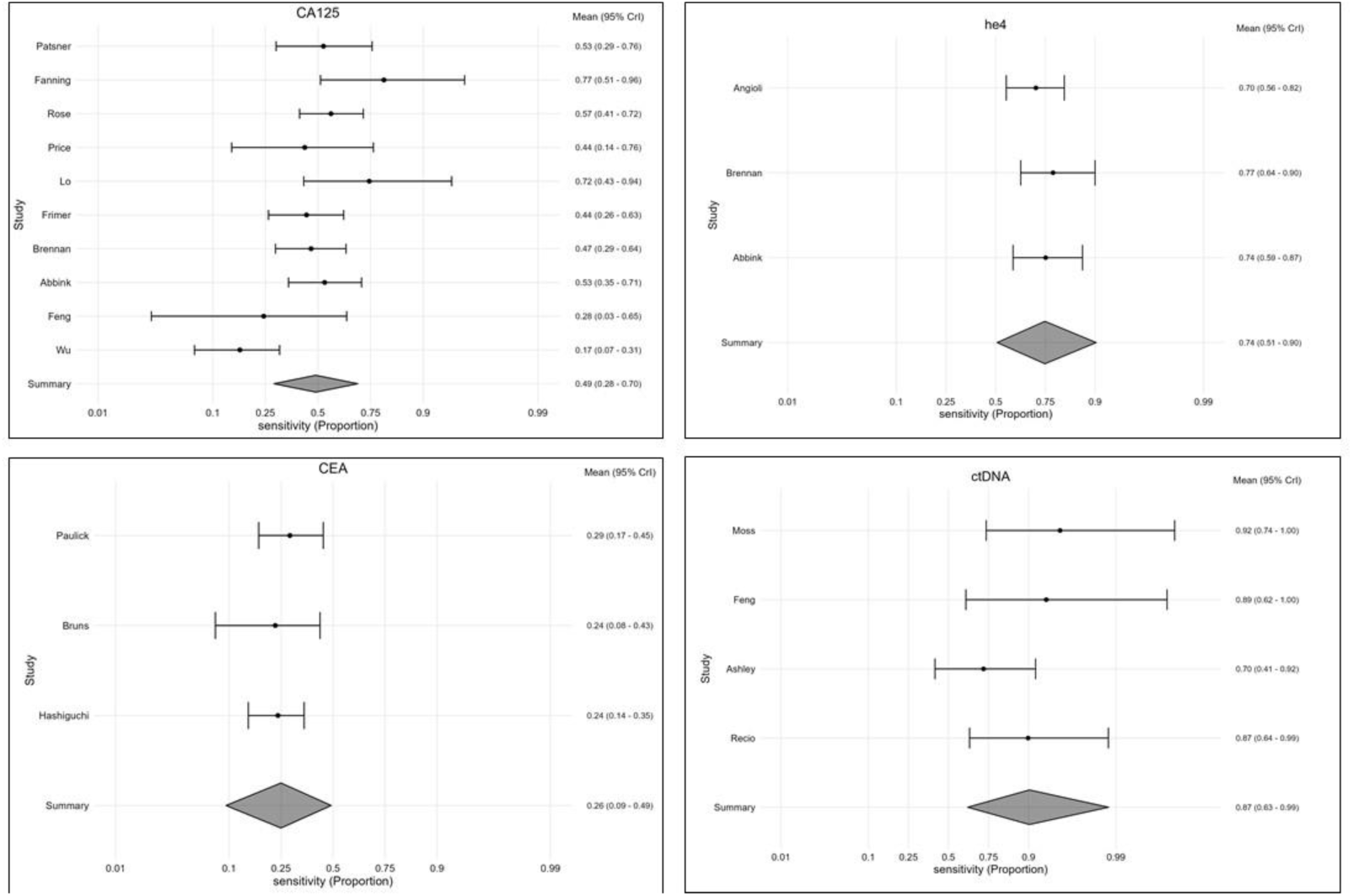
Forest plots for specificity of CA125, HE4, CEA and cf/ctDNA to detect endometrial cancer recurrence.

**Figure 3.**
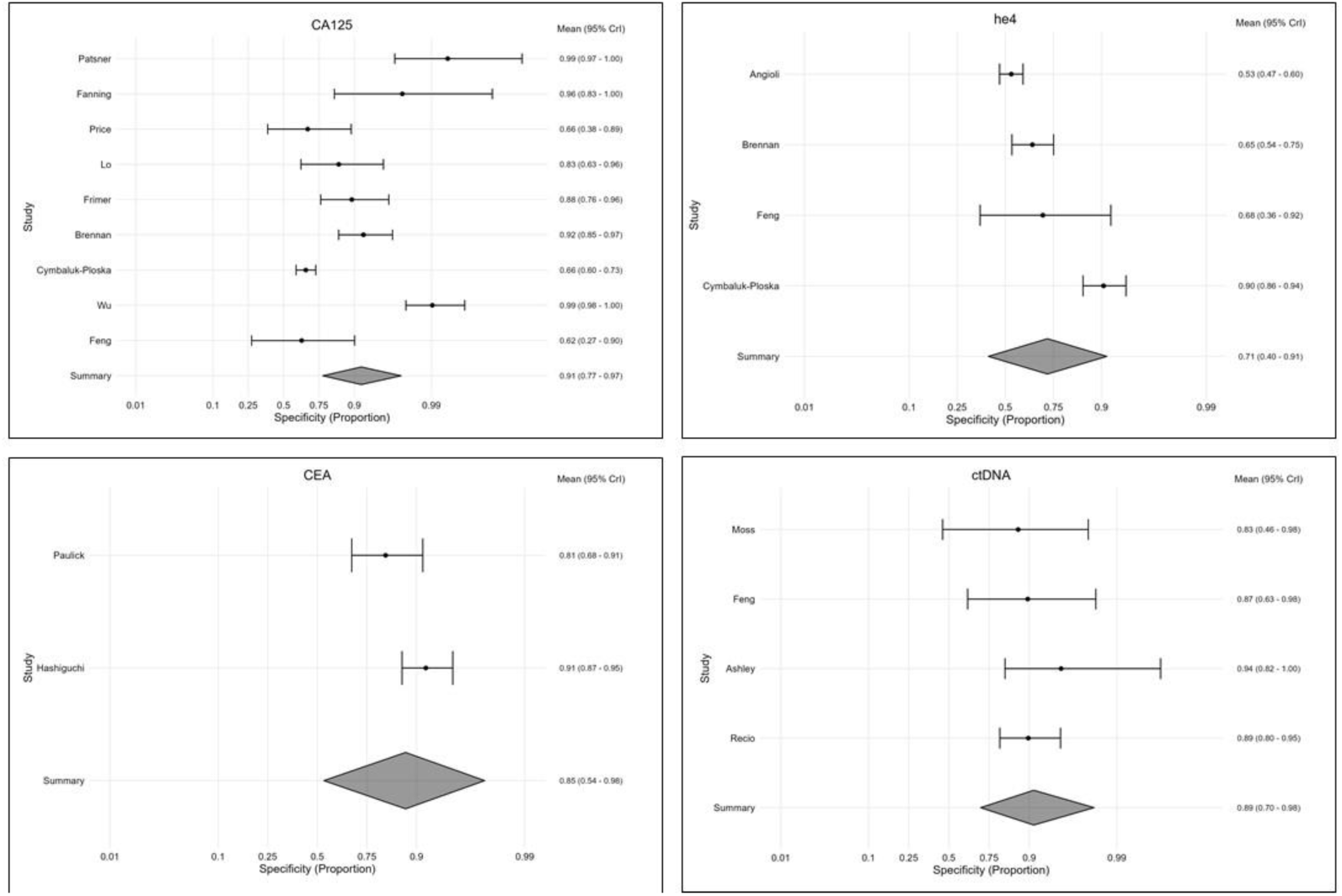
Forest plots for specificity of CA125, HE4, CEA and cf/ctDNA to detect endometrial cancer recurrence.

## DISCUSSION

This review has identified that cf/ctDNA markers have the advantage of superior sensitivity to diagnose endometrial cancer recurrence compared to the currently available blood-based biomarkers CA125 and HE4. The specificity result of cf/ctDNA may be an underestimation given that in some studies patients have had a positive ctDNA sample but not had a confirmed relapse clinically and this could have been due to the long lead time that is reported with ctDNA and the time constraints of the study rather than a true false positive result. In contrast, the specificity of CA125 also need to be interpreted with caution since bias was noted in several studies where cases with known conditions that could be associated with false positive results, for example another malignancy or infection, were excluded which could have resulted in a higher specificity rate ^(26, 27)^. The use of cf/ctDNA brings additional benefits compared to other blood -based biomarkers since it is able to provide information on the genomic profile of the recurrence and identify actionable mutations that could be used by oncologists to guide patient management, or diagnose a second malignancy ^(38)^.

Biomarkers have been successfully used in many cancer subtypes as a way of monitoring disease burden, for example the detection of biochemical prostate cancer recurrence using prostate specific antigen ^(39)^. CA125 has been traditionally known as the ovarian cancer tumour marker ^(40)^, and although high levels are associated with poor survival ^(41)^, it is not specific for ovarian cancer and has been shown to be raised in many benign gynaecological or non-gynaecological conditions ^(42)^. The routine use of CA125 as a follow-up tool to diagnose ovarian cancer recurrence and guide patient management is not standardly advocated ^(43)^ as a result of the OVO5 study which showed that CA125-guided treatment did not impact overall survival ^(44)^, however it could be argued that the management of recurrent ovarian cancer has changed radically since this trial was undertaken. CA125 and HE4 have been investigated for their ability to diagnose EC, with sensitivities of 0.71 (95% CI 0.56-0.82) and specificities of 0.87 (95% CI 0.80-0.92) for HE4 and 0.35 (95% CI 0.25-0.46) and 0.83 (95% CI 0.71-0.91) for CA125 reported ^(40)^. High pre-operative CA125 levels have also been shown to be associated with higher stage and greater risk of lymphovascular space invasion ^(45)^.

The potential of ctDNA-based biomarkers not only to diagnose but also to predict recurrence due to long lead times ^(28)^, could enable the identification of a patient population at increased risk of recurrence thereby risk stratify those who may benefit from surveillance imaging, and treatment of recurrence at the earliest opportunity. Although the psychological impact of predictive testing does need to be considered, in order not to increase anxiety and fear of cancer recurrence ^(46)^. The availability of a biomarker for endometrial cancer with high diagnostic accuracy was reported to be of interest to clinicians involved in the follow-up of endometrial cancer patients, since it was felt it would increase their confidence in starting patients on patient-initiated follow-up or introduce the possibility of a remote monitoring scheme for patients with higher-risk disease ^(47)^. The prognostic information available from ctDNA additionally has the potential to guide patient management by stratifying treatment, in particular the decision for adjuvant chemotherapy, for example the TRACC trial in colorectal cancer ^(48)^, although larger studies confirming the lead time and prognostic ability would be needed prior to such a development. Indeed the early detection of endometrial cancer recurrence at a lower tumour volume may increase the proportion of patients who would be amenable to secondary cytoreductive surgery, however identifying this population may be challenging without a sensitive/specific biomarker since imaging in asymptomatic patients has been shown to have a low detection rate ^(49)^ even in a high-risk population ^(50)^.

### Limitations

One of the key limitations of this systematic review was the heterogeneity of information presented in the publications, including histological subtype, which impacted on the number of studies that could be included in the meta-analysis. There were missing values for specificity in a number of studies, which meant the meta-analyses were performed for sensitivity and specificity separately. In addition, a number of studies reported a mean value at recurrence for all recurrent/no-recurrence cases rather than the proportion of cases with an abnormal test, which resulted in these studies being excluded from the sensitivity/specificity analysis.

## CONCLUSIONS

Integrating routine blood-based biomarker testing into endometrial cancer follow-up with a view to detect recurrence represents a transformative step in patient care. More traditional markers, including CA125 and HE4, have demonstrated some utility, however, their limitations lie in their specificity and high false positives rates. In contrast, emerging biomarkers, in particular cf/ctDNA, hold potential not only for their diagnostic ability but may have additional benefits including prognostication and genomic insights that can support more personalised treatment strategies. Given the concerning rise in endometrial cancer mortality, may be the time has come to explore the potential of a blood-based biomarker to facilitate the identification of endometrial cancer recurrence and evaluate the impact on long-term outcomes, paving the way for more effective and personalised management of endometrial cancer recurrence.

## Supplementary Materials

S1: search strategy; S2 quality assessment

## Author Contributions

Conceptualization, EM and DSG.; methodology EM, CP, SB and LW; investigation, AC, AAA, DSG and ELM; formal analysis, AC, AAA, SB, LW and ELM; writing—original draft preparation, AC, AAA and ELM; writing—review and editing, AC, CP, SB and ELM; supervision, DSG, SB and ELM. All authors have read and agreed to the published version of the manuscript.

## Institutional Review Board Statement

Not applicable

## Informed Consent Statement

Not applicable

## Data Availability Statement

Data is contained within the article or supplementary material. The original contributions presented in this study are included in the article and supplementary material. Further inquiries can be directed to the corresponding author.

## Acknowledgments

SB and LW were supported by the Leicester NIHR Biomedical Research Centre (BRC).

---

**Supplementary Data 1.** Search strategy.

| Database | Provider | No. of results |
| --- | --- | --- |
| Medline | Ovid | 3790 |
| Embase | Ovid | 5651 |
| Emcare | Ovid | 3586 |
| CINAHL | EBSCO | 442 |
| CENTRAL | Wiley | 1046 |
| OpenGrey | - | 18 |

Total: 14533

Duplicates removed: 3890

Total for screening: 10643

Limits applied to this search:

Human-only studies using part of the Cochrane RCT filters:

Cochrane RCT filters. Lefebvre C, Glanville J, Briscoe S, Featherstone R, Littlewood A, Metzendorf M-I, Noel-Storr A, Paynter R, Rader T, Thomas J, Wieland LS. Chapter 4: Searching for and selecting studies. <u>In</u>: Higgins JPT, Thomas J, Chandler J, Cumpston M, Li T, Page MJ, Welch VA (editors). Cochrane Handbook for Systematic Reviews of Interventions version 6.4 (updated October 2023). Cochrane, 2023. Available from www.training.cochrane.org/handbook

1. (endometri* adj5 (cancer* or neoplas* or carcinom* or adenocarcinom* or sarcoma* or malignan* or tumo?r* or EIN)).tw.
2. (uter* adj5 (cancer* or neoplas* or carcinom* or adenocarcinom* or sarcoma* or malignan* or tumo?r* or EIN)).tw.
3. (womb* adj5 (cancer* or neoplas* or carcinom* or adenocarcinom* or sarcoma* or malignan* or tumo?r* or EIN)).tw.
4. exp endometrial neoplasms/
5. uterine neoplasms/
6. Carcinoma, Endometrioid/
7. exp endometrial stromal tumors/
8. 1 or 2 or 3 or 4 or 5 or 6 or 7
9. ccfDNA.tw.
10. cfDNA.tw.
11. cf-NA.tw.
12. cfNA.tw.
13. ctDNA.tw.
14. cfrna.tw.
15. cirDNA.tw.
16. cirRNA.tw.
17. ((cell-free or "cell free") adj2 (serum or DNA or rna or "deoxyribonucleic acid*" or "nucleic acid*" or "ribonucleic acid*")).tw.
18. (circulat* adj3 (DNA or "nucleic acid*" or "tumo?r* DNA" or rna or "deoxyribonucleic acid*" or "ribonucleic acid*")).tw.
19. (free adj2 DNA).tw.
20. ptDNA.tw.
21. (plasma adj3 DNA).tw.
22. (serum adj (DNA or cell-free)).tw.
23. microRNA.tw.
24. miRNA.tw.
25. CA125.tw.
26. CA-125.tw.
27. cancer antigen 125.tw.
28. mucin 16.tw.
29. MUC16.tw.
30. MUC-16.tw.
31. microsatellite instability.tw.
32. MSI.tw.
33. MSI-H.tw.
34. (microsatellite adj3 unstable).tw.
35. HE4.tw.
36. HE-4.tw.
37. human epididymis 4.tw.
38. human epididymis secretory protein 4.tw.
39. serum amyloid A.tw.
40. SAA.tw.
41. SAA1.tw.
42. exp Cell-Free Nucleic Acids/
43. CA-125 Antigen/
44. Microsatellite Instability/
45. Serum Amyloid A Protein/
46. biomarker*.tw.
47. marker*.tw.
48. proteomic*.tw.
49. genomic*.tw.
50. epigenetic*.tw.
51. (("non-invasive" or "minimally invasive") adj3 (blood or serum or plasma or circulat* or test* or analys*)).tw.
52. exp Biomarkers/
53. exp Proteomics/
54. Genomics/
55. Epigenomics/
56. 9 or 10 or 11 or 12 or 13 or 14 or 15 or 16 or 17 or 18 or 19 or 20 or 21 or 22 or 23 or 24 or 25 or 26 or 27 or 28 or 29 or 30 or 31 or 32 or 33 or 34 or 35 or 36 or 37 or 38 or 39 or 40 or 41 or 42 or 43 or 44 or 45 or 46 or 47 or 48 or 49 or 50 or 51 or 52 or 53 or 54 or 55
57. exp "Sensitivity and Specificity"/
58. sensitivity.tw.
59. specificity.tw.
60. ((pre-test or pretest) adj probability).tw.
61. post-test probability.tw.
62. predictive value$.tw.
63. likelihood ratio$.tw.
64. exp endometrial neoplasms/di
65. uterine neoplasms/di
66. Carcinoma, Endometrioid/di
67. exp endometrial stromal tumors/di
68. 57 or 58 or 59 or 60 or 61 or 62 or 63 or 64 or 65 or 66 or 67
69. exp animals/ not humans.sh.
70. 8 and 56 and 68
71. 70 not 69

## Ovid Embase <1974 to 2024 November 14>

1. (endometri* adj5 (cancer* or neoplas* or carcinom* or adenocarcinom* or sarcoma* or malignan* or tumo?r* or EIN)).tw.
2. (uter* adj5 (cancer* or neoplas* or carcinom* or adenocarcinom* or sarcoma* or malignan* or tumo?r* or EIN)).tw.
3. (womb* adj5 (cancer* or neoplas* or carcinom* or adenocarcinom* or sarcoma* or malignan* or tumo?r* or EIN)).tw.
4. endometrium tumor/
5. endometrial stromal tumor/
6. exp endometrium cancer/
7. uterus tumor/
8. uterus cancer/
9. uterus carcinoma/
10. uterus sarcoma/
11. 1 or 2 or 3 or 4 or 5 or 6 or 7 or 8 or 9 or 10
12. ccfDNA.tw.
13. cfDNA.tw.
14. cf-NA.tw.
15. cfNA.tw.
16. ctDNA.tw.
17. cfrna.tw.
18. cirDNA.tw.
19. cirRNA.tw.
20. ((cell-free or "cell free") adj2 (serum or DNA or rna or "deoxyribonucleic acid*" or "nucleic acid*" or "ribonucleic acid*")).tw.
21. (circulat* adj3 (DNA or "nucleic acid*" or "tumo?r* DNA" or rna or "deoxyribonucleic acid*" or "ribonucleic acid*")).tw.
22. (free adj2 DNA).tw.
23. ptDNA.tw.
24. (plasma adj3 DNA).tw.
25. (serum adj (DNA or cell-free)).tw.
26. microRNA.tw.
27. miRNA.tw.
28. CA125.tw.
29. CA-125.tw.
30. cancer antigen 125.tw.
31. mucin 16.tw.
32. MUC16.tw.
33. MUC-16.tw.
34. microsatellite instability.tw.
35. MSI.tw.
36. MSI-H.tw.
37. (microsatellite adj3 unstable).tw.
38. HE4.tw.
39. HE-4.tw.
40. human epididymis 4.tw.
41. human epididymis secretory protein 4.tw.
42. WAP four-disulfide core domain protein 2/
43. serum amyloid A.tw.
44. SAA.tw.
45. SAA1.tw.
46. exp cell free nucleic acid/
47. CA 125 antigen/
48. microsatellite instability/
49. serum amyloid A/
50. biomarker*.tw.
51. marker*.tw.
52. proteomic*.tw.
53. genomic*.tw.
54. epigenetic*.tw.
55. (("non-invasive" or "minimally invasive") adj3 (blood or serum or plasma or circulat* or test* or analys*)).tw.
56. biological marker/
57. exp proteomics/
58. genomics/
59. epigenetics/
60. 12 or 13 or 14 or 15 or 16 or 17 or 18 or 19 or 20 or 21 or 22 or 23 or 24 or 25 or 26 or 27 or 28 or 29 or 30 or 31 or 32 or 33 or 34 or 35 or 36 or 37 or 38 or 39 or 40 or 41 or 42 or 43 or 44 or 45 or 46 or 47 or 48 or 49 or 50 or 51 or 52 or 53 or 54 or 55 or 56 or 57 or 58 or 59
61. exp "SENSITIVITY AND SPECIFICITY"/
62. sensitivity.tw.
63. specificity.tw.
64. ((pre-test or pretest) adj probability).tw.
65. post-test probability.tw.
66. predictive value$.tw.
67. likelihood ratio$.tw.
68. *Diagnostic Accuracy/
69. endometrium tumor/di
70. endometrial stromal tumor/di
71. exp endometrium cancer/di
72. uterus tumor/di
73. uterus cancer/di
74. uterus carcinoma/di
75. uterus sarcoma/di
76. 61 or 62 or 63 or 64 or 65 or 66 or 67 or 68 or 69 or 70 or 71 or 72 or 73 or 74 or 75
77. exp experimental organism/
78. animal tissue/
79. animal cell/
80. exp animal disease/
81. exp carnivore disease/
82. exp bird/
83. exp experimental animal welfare/
84. exp animal husbandry/
85. animal behavior/
86. exp animal cell culture/
87. exp mammalian disease/
88. exp mammal/
89. exp marine species/
90. nonhuman/
91. animal.hw.
92. 77 or 78 or 79 or 80 or 81 or 82 or 83 or 84 or 85 or 86 or 87 or 88 or 89 or 90 or 91
93. 92 not human/
94. 11 and 60 and 76
95. 94 not 93

## Ovid Emcare <1995 to 2024 Week 45>

1. (endometri* adj5 (cancer* or neoplas* or carcinom* or adenocarcinom* or sarcoma* or malignan* or tumo?r* or EIN)).tw.
2. (uter* adj5 (cancer* or neoplas* or carcinom* or adenocarcinom* or sarcoma* or malignan* or tumo?r* or EIN)).tw.
3. (womb* adj5 (cancer* or neoplas* or carcinom* or adenocarcinom* or sarcoma* or malignan* or tumo?r* or EIN)).tw.
4. endometrium tumor/
5. endometrial stromal tumor/
6. exp endometrium cancer/
7. uterus tumor/
8. uterus cancer/
9. uterus carcinoma/
10. uterus sarcoma/
11. 1 or 2 or 3 or 4 or 5 or 6 or 7 or 8 or 9 or 10
12. ccfDNA.tw.
13. cfDNA.tw.
14. cf-NA.tw.
15. cfNA.tw.
16. ctDNA.tw.
17. cfrna.tw.
18. cirDNA.tw.
19. cirRNA.tw.
20. ((cell-free or "cell free") adj2 (serum or DNA or rna or "deoxyribonucleic acid*" or "nucleic acid*" or "ribonucleic acid*")).tw.
21. (circulat* adj3 (DNA or "nucleic acid*" or "tumo?r* DNA" or rna or "deoxyribonucleic acid*" or "ribonucleic acid*")).tw.
22. (free adj2 DNA).tw.
23. ptDNA.tw.
24. (plasma adj3 DNA).tw.
25. (serum adj (DNA or cell-free)).tw.
26. microRNA.tw.
27. miRNA.tw.
28. CA125.tw.
29. CA-125.tw.
30. cancer antigen 125.tw.
31. mucin 16.tw.
32. MUC16.tw.
33. MUC-16.tw.
34. microsatellite instability.tw.
35. MSI.tw.
36. MSI-H.tw.
37. (microsatellite adj3 unstable).tw.
38. HE4.tw.
39. HE-4.tw.
40. human epididymis 4.tw.
41. human epididymis secretory protein 4.tw.
42. WAP four-disulfide core domain protein 2/
43. serum amyloid A.tw.
44. SAA.tw.
45. SAA1.tw.
46. exp cell free nucleic acid/
47. CA 125 antigen/
48. microsatellite instability/
49. serum amyloid A/
50. biomarker*.tw.
51. marker*.tw.
52. proteomic*.tw.
53. genomic*.tw.
54. epigenetic*.tw.
55. (("non-invasive" or "minimally invasive") adj3 (blood or serum or plasma or circulat* or test* or analys*)).tw.
56. biological marker/
57. exp proteomics/
58. genomics/
59. epigenetics/
60. 12 or 13 or 14 or 15 or 16 or 17 or 18 or 19 or 20 or 21 or 22 or 23 or 24 or 25 or 26 or 27 or 28 or 29 or 30 or 31 or 32 or 33 or 34 or 35 or 36 or 37 or 38 or 39 or 40 or 41 or 42 or 43 or 44 or 45 or 46 or 47 or 48 or 49 or 50 or 51 or 52 or 53 or 54 or 55 or 56 or 57 or 58 or 59
61. exp experimental organism/
62. animal tissue/
63. animal cell/
64. exp animal disease/
65. exp carnivore disease/
66. exp bird/
67. exp experimental animal welfare/
68. exp animal husbandry/
69. animal behavior/
70. exp animal cell culture/
71. exp mammalian disease/
72. exp mammal/
73. exp marine species/
74. nonhuman/
75. animal.hw.
76. 61 or 62 or 63 or 64 or 65 or 66 or 67 or 68 or 69 or 70 or 71 or 72 or 73 or 74 or 75 7319180
77. 76 not human/
78. 11 and 60
79. 78 not 77

## Cochrane Central Register of Controlled Trials – CENTRAL

#1 endometri* NEAR/5 (cancer* or neoplas* or carcinom* or adenocarcinom* or sarcoma* or malignan* or tumo?r* or EIN)
#2 uter* NEAR/5 (cancer* or neoplas* or carcinom* or adenocarcinom* or sarcoma* or malignan* or tumo?r* or EIN)
#3 womb* NEAR/5 (cancer* or neoplas* or carcinom* or adenocarcinom* or sarcoma* or malignan* or tumo?r* or EIN)
#4 MeSH descriptor: [Endometrial Neoplasms] 3 tree(s) exploded #5 MeSH descriptor: [Uterine Neoplasms] this term only
#6 MeSH descriptor: [Carcinoma, Endometrioid] this term only
#7 MeSH descriptor: [Endometrial Stromal Tumors] 4 tree(s) exploded #8 {OR #1-#7}
#9 ccfDNA
#10 cfDNA
#11 "cf-NA"
#12 cfNA
#13 ctDNA
#14 cfrna
#15 cirDNA
#16 cirRNA
#17 (cell-free or "cell free") NEAR/2 (serum or DNA or rna or (deoxyribonucleic NEXT acid*) or (nucleic NEXT acid*) or (ribonucleic NEXT acid*))
#18 circulat* NEAR/3 (DNA or (nucleic NEXT acid*) or (tumo?r* NEXT DNA) or rna or (deoxyribonucleic NEXT acid*) or (ribonucleic NEXT acid*))<colcnt=2>
#19 free NEAR/2 DNA
#20 ptDNA
#21 plasma NEAR/3 DNA
#22 serum NEXT (DNA or cell-free)
#23 microRNA
#24 miRNA
#25 CA125
#26 "CA-125"
#27 "cancer antigen 125"
#28 "mucin 16"
#29 MUC16
#30 "MUC-16"
#31 "microsatellite instability"
#32 MSI
#33 "MSI-H"
#34 microsatellite NEAR/3 unstable
#35 HE4
#36 "HE-4"
#37 "human epididymis 4"
#38 "human epididymis secretory protein 4"
#39 "serum amyloid A" #40 SAA
#41 SAA1
#42 MeSH descriptor: [Cell-Free Nucleic Acids] explode all trees
#43 MeSH descriptor: [CA-125 Antigen] this term only
#44 MeSH descriptor: [Microsatellite Instability] this term only #45 MeSH descriptor: [Serum Amyloid A Protein] this term only
#46 biomarker*
#47 marker*
#48 proteomic*
#49 genomic*
#50 epigenetic*
#51 ("non-invasive" or "minimally invasive") NEAR/3 (blood or serum or plasma or circulat* or test* or analys*)
#52 MeSH descriptor: [Biomarkers] explode all trees
#53 MeSH descriptor: [Proteomics] explode all trees
#54 MeSH descriptor: [Genome] explode all trees
#55 MeSH descriptor: [Epigenomics] explode all trees
#56 {OR #9-#55}
#57 #8 AND #56

## CINAHL (EBSCO) – 15^th^ November 2024

S1 TI ( (endometri* N5 (cancer* or neoplas* or carcinom* or adenocarcinom* or sarcoma* or malignan* or tumo?r* or EIN)) ) OR AB ( (endometri* N5 (cancer* or neoplas* or carcinom* or adenocarcinom* or sarcoma* or malignan* or tumo?r* or EIN)) )
S2 TI ( (uter* N5 (cancer* or neoplas* or carcinom* or adenocarcinom* or sarcoma* or malignan* or tumo?r* or EIN)) ) OR AB ( (uter* N5 (cancer* or neoplas* or carcinom* or adenocarcinom* or sarcoma* or malignan* or tumo?r* or EIN)) )
S3 TI ( (womb* N5 (cancer* or neoplas* or carcinom* or adenocarcinom* or sarcoma* or malignan* or tumo?r* or EIN)) ) OR AB ( (womb* N5 (cancer* or neoplas* or carcinom* or adenocarcinom* or sarcoma* or malignan* or tumo?r* or EIN)) )
S4 (MH "Endometrial Neoplasms")
S5 (MH "Uterine Neoplasms")
S6 S1 OR S2 OR S3 OR S4 OR S5
S7 TI ccfDNA OR AB ccfDNA
S8 TI cfDNA OR AB cfDNA
S9 TI cf-NA OR AB cf-NA
S10 TI cfNA OR AB cfNA
S11 TI ctDNA OR AB ctDNA
S12 TI cfrna OR AB cfrna
S13 TI cirDNA OR AB cirDNA
S14 TI cirRNA OR AB cirRNA
S15 TI ( (cell-free or "cell free") N2 (serum or DNA or rna or "deoxyribonucleic acid*" or "nucleic acid*" or "ribonucleic acid*") ) OR AB ( (cell-free or "cell free") N2 (serum or DNA or rna or "deoxyribonucleic acid*" or "nucleic acid*" or "ribonucleic acid*") )
S16 TI ( circulat* N3 (DNA or "nucleic acid*" or "tumo? r* DNA" or rna or "deoxyribonucleic acid*" or "ribonucleic acid*") ) OR AB ( circulat* N3 (DNA or "nucleic acid*" or "tumo?r* DNA" or rna or "deoxyribonucleic acid*" or "ribonucleic acid*") )
S17 TI free N2 DNA OR AB free N2 DNA
S18 TI ptDNA OR AB ptDNA
S19 TI plasma N3 DNA OR AB plasma N3 DNA
S20 TI ( serum N1 (DNA or cell-free) ) OR AB ( serum N1 (DNA or cellfree) )
S21 TI microRNA OR AB microRNA
S22 TI miRNA OR AB miRNA S23 TI CA125 OR AB CA125 S24 TI CA-125 OR AB CA125
S25 TI "cancer antigen 125" OR AB "cancer antigen 125" S26 TI "mucin 16" OR AB "mucin 16"
S27 TI MUC16 OR AB MUC16
S28 TI MUC-16 OR AB MUC16
S29 TI "microsatellite instability" OR AB "microsatellite instability"
S30 TI MSI OR AB MSI
S31 TI MSI-H OR AB MSI-H
S32 TI microsatellite N3 unstable OR AB microsatellite N3 unstable
S33 TI HE4 OR AB HE4
S34 TI HE-4 OR AB HE-4
S35 TI "human epididymis 4" OR AB "human epididymis 4"
S36 TI "human epididymis secretory protein 4" OR AB "human epididymis secretory protein 4"
S37 TI "serum amyloid A" OR AB "serum amyloid A" S38 TI SAA OR AB SAA
S39 TI SAA1 OR AB SAA1
S40 (MH "Cell-Free Nucleic Acids") S41 TI biomarker* OR AB biomarker* S42 TI marker* OR AB marker*
S43 TI proteomic* OR AB proteomic* S44 TI genomic* OR AB genomic* S45 TI epigenetic* OR AB epigenetic*
S46 TI ( ("non-invasive" or "minimally invasive") N3 (blood or serum or plasma or circulat* or test* or analys*) ) OR AB ( ("non-invasive" or "minimally invasive") N3 (blood or serum or plasma or circulat* or test* or analys*) )
S47 (MH "Biological Markers")
S48 (MH "Tumor Markers, Biological+")
S49 (MH "Proteomics+")
S50 (MH "Genomics")
S51 (MH "Epigenomics")
S52 S7 OR S8 OR S9 OR S10 OR S11 OR S12 OR S13 OR S14 OR S15 OR S16 OR S17 OR S18 OR S19 OR S20 OR S21 OR S22 OR S23 OR S24 OR S25 OR S26 OR S27 OR S28 OR S29 OR S30 OR S31 OR S32 OR S33 OR S34 OR S35 OR S36 OR S37 OR S38 OR S39 OR S40 OR S41 OR S42 OR S43 OR S44 OR S45 OR S46 OR S47 OR S48 OR S49 OR S50 OR S51
S53 (MH "Sensitivity and Specificity")
S54 TI sensitivity OR AB sensitivity
S55 TI specificity OR AB specificity
S56 TI ( (pre-test or pretest) N1 probability ) OR AB ( (pre-test or pretest) N1 probability )
S57 TI "post-test probability" OR AB "post-test probability"
S58 TI "predictive value*" OR AB "predictive value*"
S59 TI "likelihood ratio*" OR AB "likelihood ratio*"
S60 (MH "Endometrial Neoplasms/DI")
S61 (MH "Uterine Neoplasms/DI")
S62 S53 OR S54 OR S55 OR S56 OR S57 OR S58 OR S59 OR S60 OR S61
S63 S6 AND S52 AND S62

## penGrey strategy

((endometri* OR uter* OR womb*) NEAR/5 (cancer* OR neoplas* OR carcinom* OR adenocarcinom* OR sarcoma* OR malignan* OR tumor* OR tumour* OR EIN)) AND (DNA OR RNA OR "deoxyribonucleic acid" OR "nucleic acid" OR "ribonucleic acid" OR CA125 OR "CA-125" OR "cancer antigen 125" OR "mucin 16" OR MUC16 OR "MUC-16" OR "microsatellite instability" OR MSI OR "MSI-H" OR (microsatellite NEAR/3 unstable) OR HE4 OR "HE-4" OR "human epididymis 4" OR "human epididymis secretory protein 4" OR "serum amyloid A" OR SAA OR SAA1 OR biomarker* OR marker* OR proteomic* OR genomics* OR epigenetic*)

**18 results on 3^rd^ June 2020 (has since been discontinued)**

**Supplementary data 2.**
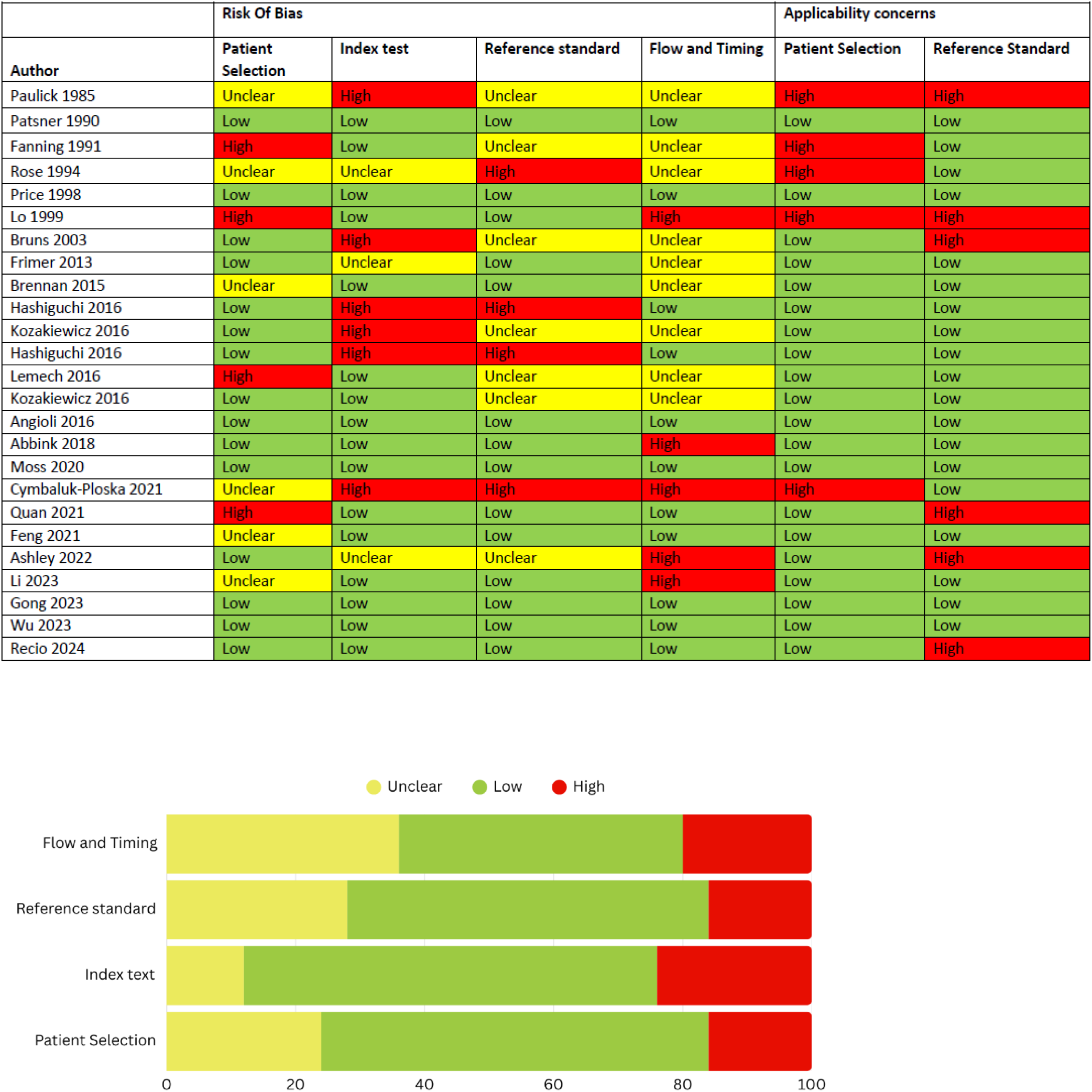
Quality assessment (QUADAS 2) of included studies.

## REFERENCES

[1] Cancer Research UK. Uterine cancer mortality statistics. 2023.

[2] Lees B, Hampton JM, Trentham-Dietz A et al. A population-based study of causes of death after endometrial cancer according to major risk factors. Gynecol Oncol. 2021;160: 655–9.

[3] Rashid T, Bennett JE, Muller DC et al. Mortality from leading cancers in districts of England from 2002 to 2019: a population-based, spatiotemporal study. Lancet Oncol. 2024;25: 86–98.

[4] Moss EL, Teece L, Darko N. Uterine cancer mortality and Black women: time to act. Lancet Oncol. 2023;24: 586–8.

[5] Harter P, Sehouli J, Vergote I et al. Randomized Trial of Cytoreductive Surgery for Relapsed Ovarian Cancer. N Engl J Med. 2021;385: 2123–31.

[6] Ren Y, Shan B, Shi D, Wang H. Salvage cytoreductive surgery for patients with recurrent endometrial cancer: a retrospective study. BMC Cancer. 2014;14: 135.

[7] Robbins JR, Yechieli R, Laser B et al. Is time to recurrence after hysterectomy predictive of survival in patients with early stage endometrial carcinoma? Gynecol Oncol. 2012;127: 38–42.

[8] Boruta DM, 2nd, Gehrig PA, Fader AN, Olawaiye AB. Management of women with uterine papillary serous cancer: a Society of Gynecologic Oncology (SGO) review. Gynecol Oncol. 2009;115: 142–53.

[9] Zola P, Ciccone G, Piovano E et al. Effectiveness of Intensive Versus Minimalist Follow-Up Regimen on Survival in Patients With Endometrial Cancer (TOTEM Study): A Randomized, Pragmatic, Parallel Group, Multicenter Trial. J Clin Oncol. 2022;40: 3817–27.

[10] Riedinger CJ, Patterson JM, Backes FJ et al. The contemporary presentation and diagnosis of endometrial cancer recurrence: When, where, and how? Gynecol Oncol. 2022;167: 174–80.

[11] Kommoss S, McConechy MK, Kommoss F et al. Final validation of the ProMisE molecular classifier for endometrial carcinoma in a large population-based case series. Ann Oncol. 2018;29: 1180–8.

[12] Quan Q, Liao Q, Yin W et al. Serum HE4 and CA125 combined to predict and monitor recurrence of type II endometrial carcinoma. Sci Rep. 2021;11: 21694.

[13] Wan JCM, Mughal TI, Razavi P et al. Liquid biopsies for residual disease and recurrence. Med. 2021;2: 1292–313.

[14] Patsner B, Orr JW, Jr., Mann WJ, Jr. Use of serum CA 125 measurement in posttreatment surveillance of early-stage endometrial carcinoma. Am J Obstet Gynecol. 1990;162: 427–9.

[15] Fanning J, Piver MS. Serial CA 125 levels during chemotherapy for metastatic or recurrent endometrial cancer. Obstet Gynecol. 1991;77: 278–80.

[16] Rose PG, Sommers RM, Reale FR et al. Serial serum CA 125 measurements for evaluation of recurrence in patients with endometrial carcinoma. Obstet Gynecol. 1994;84: 12–6.

[17] Price FV, Chambers SK, Carcangiu ML et al. CA 125 may not reflect disease status in patients with uterine serous carcinoma. Cancer. 1998;82: 1720–5.

[18] Lo SS, Khoo US, Cheng DK et al. Role of serial tumor markers in the surveillance for recurrence in endometrial cancer. Cancer Detect Prev. 1999;23: 397–400.

[19] Frimer M, Hou JY, McAndrew TC et al. The clinical relevance of rising CA-125 levels within the normal range in patients with uterine papillary serous cancer. Reprod Sci. 2013;20: 449–55.

[20] Gong S, Quan Q, Meng Y et al. The value of serum HE4 and CA125 levels for monitoring the recurrence and risk stratification of endometrial endometrioid carcinoma. Heliyon. 2023;9: e18016.

[21] Angioli R, Capriglione S, Scaletta G et al. The role of HE4 in endometrial cancer recurrence: how to choose the optimal follow-up program. Tumour Biol. 2016;37: 4973–8.

[22] Brennan DJ, Hackethal A, Mann KP et al. Serum HE4 detects recurrent endometrial cancer in patients undergoing routine clinical surveillance. BMC Cancer. 2015;15: 33.

[23] Abbink K, Zusterzeel PL, Geurts-Moespot AJ et al. HE4 is superior to CA125 in the detection of recurrent disease in high-risk endometrial cancer patients. Tumour Biol. 2018;40: 1010428318757103.

[24] Cymbaluk-Ploska A, Gargulinska P, Bulsa M et al. Can the Determination of HE4 and CA125 Markers Affect the Treatment of Patients with Endometrial Cancer? Diagnostics (Basel). 2021;11.

[25] Feng W, Jia N, Jiao H et al. Circulating tumor DNA as a prognostic marker in high-risk endometrial cancer. J Transl Med. 2021;19: 51.

[26] Li Y, Hou X, Chen W et al. Development and validation of a nomogram for predicting recurrence-free survival in endometrial cancer: a multicenter study. Sci Rep. 2023;13: 20270.

[27] Wu HH, Chou HT, Tseng JY et al. The relationship between serum CA-125 level and recurrence in surgical stage I endometrial cancer patients. J Chin Med Assoc. 2023;86: 1001–7.

[28] Moss EL, Gorsia DN, Collins A et al. Utility of Circulating Tumor DNA for Detection and Monitoring of Endometrial Cancer Recurrence and Progression. Cancers (Basel). 2020;12.

[29] Ashley CW, Selenica P, Patel J et al. High-Sensitivity Mutation Analysis of Cell-Free DNA for Disease Monitoring in Endometrial Cancer. Clin Cancer Res. 2023;29: 410–21.

[30] Recio F, Scalise CB, Loar P et al. Post-surgical ctDNA-based molecular residual disease detection in patients with stage I uterine malignancies. Gynecol Oncol. 2024;182: 63–9.

[31] Paulick R, Caffier H, Horner G et al. Clinical significance of different serum tumor markers in gynecological malignancies. Cancer Detect Prev. 1985;8: 115–20.

[32] Bruns F, Micke O, Halek G et al. Carcinoembryonic antigen (CEA)--a useful marker for the detection of recurrent disease in endometrial carcinoma patients. Anticancer Res. 2003;23: 1103–6.

[33] Hashiguchi Y, Kasai M, Fukuda T et al. Serum carcinoembryonic antigen as a tumour marker in patients with endometrial cancer. Curr Oncol. 2016;23: e439–e42.

[34] Kozakiewicz B, Chadzynska M, Dmoch-Gajzlerska E, Stefaniak M. Monitoring the treatment outcome in endometrial cancer patients by CEA and TATI. Tumour Biol. 2016;37: 9367–74.

[35] Hashiguchi Y, Kasai M, Fukuda T et al. Serum Sialyl-Tn (STN) as a Tumor Marker in Patients with Endometrial Cancer. Pathol Oncol Res. 2016;22: 501–4.

[36] Kozakiewicz B, Chadzynska M, Dmoch-Gajzlerska E, Stefaniak M. Tumor-associated trypsin inhibitor in patients with endometrial cancer. Tumori. 2016;102: 527–32.

[37] Lemech CR, Ensell L, Paterson JC et al. Enumeration and Molecular Characterisation of Circulating Tumour Cells in Endometrial Cancer. Oncology. 2016;91: 48–54.

[38] Wadsley M, Guttery DG, Cowley C et al. Design and validation of a custom circulating tumour DNA assay to detect endometrial cancer recurrence. 2025.

[39] Van den Broeck T, van den Bergh RCN, Arfi N et al. Prognostic Value of Biochemical Recurrence Following Treatment with Curative Intent for Prostate Cancer: A Systematic Review. Eur Urol. 2019;75: 967–87.

[40] Canney PA, Moore M, Wilkinson PM, James RD. Ovarian cancer antigen CA125: a prospective clinical assessment of its role as a tumour marker. Br J Cancer. 1984;50: 765–9.

[41] Wang Q, Feng X, Liu X, Zhu S. Prognostic Value of Elevated Pre-treatment Serum CA-125 in Epithelial Ovarian Cancer: A Meta-Analysis. Front Oncol. 2022;12: 868061.

[42] Moss EL, Hollingworth J, Reynolds TM. The role of CA125 in clinical practice. J Clin Pathol. 2005;58: 308–12.

[43] Moss E, Taylor A, Andreou A et al. British Gynaecological Cancer Society (BGCS) ovarian, tubal and primary peritoneal cancer guidelines: Recommendations for practice update 2024. Eur J Obstet Gynecol Reprod Biol. 2024;300: 69–123.

[44] Rustin GJ, van der Burg ME, Griffin CL et al. Early versus delayed treatment of relapsed ovarian cancer (MRC OV05/EORTC 55955): a randomised trial. Lancet. 2010;376: 1155–63.

[45] Shawn LyBarger K, Miller HA, Frieboes HB. CA125 as a predictor of endometrial cancer lymphovascular space invasion and lymph node metastasis for risk stratification in the preoperative setting. Sci Rep. 2022;12: 19783.

[46] Relton A, Collins A, Guttery DS et al. Patient acceptability of circulating tumour DNA testing in endometrial cancer follow-up. Eur J Cancer Care (Engl). 2021;30: e13429.

[47] Amirthanayagam A, Boulter L, Millet N et al. Risk Stratified Follow-Up for Endometrial Cancer: The Clinicians’ Perspective. Curr Oncol. 2023;30: 2237–48.

[48] Slater S, Bryant A, Chen HC et al. ctDNA guided adjuvant chemotherapy versus standard of care adjuvant chemotherapy after curative surgery in patients with high risk stage II or stage III colorectal cancer: a multi-centre, prospective, randomised control trial (TRACC Part C). BMC Cancer. 2023;23: 257.

[49] Fung-Kee-Fung M, Dodge J, Elit L et al. Follow-up after primary therapy for endometrial cancer: a systematic review. Gynecol Oncol. 2006;101: 520–9.

[50] Hunn J, Tenney ME, Tergas AI et al. Patterns and utility of routine surveillance in high grade endometrial cancer. Gynecol Oncol. 2015;137: 485–9.

